# A protocol for the exploration of strategies to reduce the burden of prostate cancer in South Africa’s Eastern Cape province: A multi-methods study

**DOI:** 10.1101/2025.04.02.25325151

**Authors:** Xolelwa Ntlongweni, Sibusiso C. Nomatshila, Wezile W. Chitha, Sikhumbuzo A. Mabunda

## Abstract

Prostate cancer is the second most diagnosed cancer in men, disproportionately affects African populations with rural South Africa, especially the Eastern Cape, facing high prevalence due to healthcare inequalities, late diagnoses and limited resources. This study will play a role in aiding the responsiveness of the Eastern Cape Department of Health to deliver optimal cancer prevention and control for prostate cancer. Furthermore, it will highlight ways to destigmatise prostate cancer in African communities.

**Methods:** A multi-methods approach conducted in the OR Tambo district, Eastern Cape. Phase 1 involves a scoping review to explore prostate cancer prevention strategies in Africa. Phase 2 is a quantitative cross-sectional study analysing secondary data of prostate cancer survivors. Phase 3 and 4 include qualitative interviews to assess survivors’ perceptions and the general male population’s knowledge and attitudes towards prostate cancer care and attitudes towards prophylactic orchidectomy.

**Analysis:** Phase 1, narrative synthesis of articles, summarising findings with tables, a PRISMA diagram for article selection and exclusions. Phase 2; data will be analysed using descriptive statistics, T-tests, ANOVA, logistic regression and chi-square tests in STATA. Phase 3; thematic analysis of semi-structured interviews using NVIVO 14 using inductive coding and a six-step approach to identify key themes. Phase 4; analysed using NVIVO 14 to identify themes related to PC knowledge and health behaviours.

**Ethics and Dissemination:** This study received ethics approval from the Walter Sisulu University Health Sciences Research Ethics Committee with Ethics approval number WSUHREC014/2025 Informed written consent will be obtained for primary data, while secondary data will be anonymised. Findings will be shared through publications and conferences to advance prostate cancer prevention.

## Introduction

Cancer remains a global burden with 14.1 million new cases and 8.2 million deaths reported in 2012, a figure projected to grow significantly by 2030 [1,2]. Similarly, prostate cancer is a significant health concern, being the second most diagnosed cancer in men and the fifth leading cause of cancer-related deaths globally, with an estimated 1.6 million diagnoses and 366,000 deaths annually [3,4,5]. The incidence of prostate cancer is particularly high in Africa, with prevalence rates significantly exceeding global averages, especially in South Africa [6,7].

African men are disproportionately affected by prostate cancer due to factors like genetics, poverty and limited access to healthcare services. The disease progresses more aggressively and is often diagnosed late in black men, particularly in rural areas [8,2]. Despite the presence of cancer registries like the one in South Africa’s Eastern Cape province, underreporting remains a challenge due to the exclusion of non-biopsy-confirmed cases [9].

South Africa’s health care inequalities exacerbate the problem. Cancer care resources, including oncology staff, are concentrated in urban provinces, leaving rural regions like the Eastern Cape underserved. For instance, the province lacks adequate oncology personnel, impacting survival and quality of life for prostate cancer patients [10.11]. Additionally, South African black males are less likely to seek formal healthcare and often consult traditional healers first [12].

Cancer care services are distributed unequally between South Africa’s rural and urban provinces [11]. Service delivery is concentrated in the urban Western Cape and Gauteng provinces. Provinces such as the Eastern Cape, Limpopo, Mpumalanga, Free State and KwaZulu-Natal have limited access to diagnostic, curative, rehabilitative, psychosocial or palliative care services for cancer [11].

Furthermore, lack of awareness among the general public, and limited awareness and/or competencies among staff has negative impacts on the prognosis of patients with cancer [11]. This either results in delayed or undiagnosed cancers and at times results in delayed referral to the appropriate services [11].

With the increasing demand of cancer care services, it is therefore important that treatment centres are staffed and equipped in-order to manage this demand. It is therefore necessary to also determine the barriers linked to staff retention for cancer care services in the Eastern Cape. Because this study is in the Eastern Cape province, it may be important to look at norms and values of men in the rural areas of the province.

Prostate cancer is a problem that is inclusive of all stakeholders and their collaboration is key in combating its devastating effects. In 2012 the incidence of prostate cancer was 20.5 per 100 000 and is the leading cancer observed among men [11].

Lusikisiki (a region that refers patients to Nelson Mandela Academic Hospital) recorded an incidence (53.1 per 100 000) that was almost two times high than the global incidence (29.3 per 100 000) [13] in the period between 2013 to 2017.

There is a need for implementing more accurate strategies for prevention of prostate cancer, the recommendations of this study will be tailored for the Eastern Cape community depending on the disease profile and the province’s specific needs. Information from this study will also play a role in planning for health, cancer care policies and the allocation of resources.

## Objectives

Determine socio-demographic characteristics of prostate cancer patients.

Identify social norms, values, experiences, and perceptions of men related to prostate cancer prevention in OR Tambo, Eastern Cape province of South Africa.

Explore the gaps between men’s health behaviour and men’s health-seeking behaviour in OR Tambo region, Eastern Cape province of South Africa.

Develop health behavioral strategies to reduce prostate cancer burden in OR Tambo region, Eastern Cape province of South Africa.

## Materials and methods

### Study Setting

This study will be conducted in the OR Tambo district in the Eastern Cape Province, in South Africa.

The Eastern Cape Province has eight (8) health districts, OR Tambo has a population of 1,760,389, making it the biggest district in the Eastern Cape Province [14].

OR Tambo district has five sub-districts as illustrated in Figure 1, namely; King Sabata Dalindyebo sub district (KSD), Mhlontlo sub-district, Nyandeni sub district, Port St Johns and Ngquza Hill sub district^14^. Of the total population, 9.3% lives in urban areas whereas the remaining population lives in a largely rural area [14].

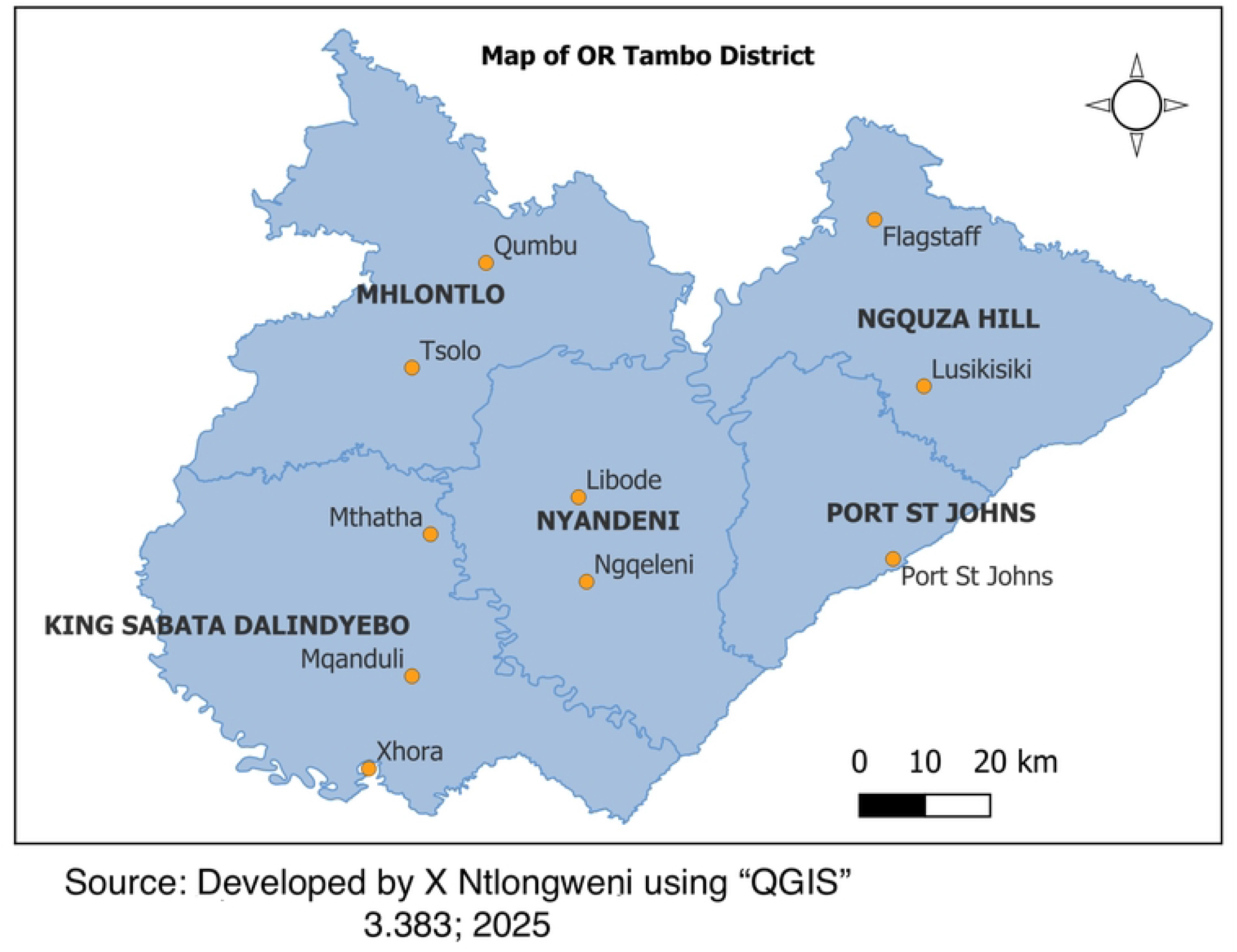

The entire OR Tambo District has two regional hospitals, 12 district hospitals, 11 community health centres, 49 clinics, 52 health posts and 61 mobiles. All the facilities refer cancer patients to the one Central hospital (Nelson Mandela Academic Hospital [14].

According to the Eastern Cape Cancer Incidence Report 2013-2017 the incidence of prostate cancer is highest in Lusikisiki (53.1%), and Flagstaff (14.1%), this constitutes 88% of the total prostate cancer cases to originate from the OR Tambo District. This is why this study will be conducted in this district.

### Study Design

The study’s objectives will be answered through a multi-methods approach of four sub-studies. Fig 1 OR Tambo District is located in the Eastern Cape, created by author (Ntlongweni, 2025).

### Phase 1: Scoping review

A scoping review will be conducted to explore strategies that have been used to reduce prostate cancer in Africa. The scoping review will further aid the identification of existing or emerging body of evidence and to determine possible gaps on the epidemiology of prostate cancer and effective strategies that have been used for prevention in a rural African context. The framework used for the review is based on the one set out by Arksey and O’Malley, 2005 [15]. The process will be documented and will provide plenty of detail, this is to increase the reliability of findings and ensure that there is undeniable methodological rigor. The approach will use the six stages of a scoping review; i) Identifying the research question, ii) Identifying relevant studies, iii) Study selection, iv) Charting the data, v) Collating, summarising, and reporting results, vi) Consultation.

### Phase 2: Quantitative Cross-sectional study

A quantitative cross-sectional study that will focus on the epidemiology of prostate cancer will be undertaken among prostate cancer survivors. This sub-study will make use of secondary data of patients who were seen at Nelson Mandela Academic hospital between March 2020 and November 2021.

### Phase 3: Qualitative, Semi-structured in-depth interviews

Qualitative, semi-structured in-depth interviews of prostate cancer survivors will be undertaken to ascertain their perceptions and beliefs on prostate cancer.

### Phase 4: Qualitative, Semi-structured in-depth interviews

Qualitative, semi-structured, in-depth interviews will be undertaken among a general population of males to establish their knowledge on prostate cancer and their health seeking behaviour. This sub-study will also establish their attitudes towards orchidectomy as a strategy for managing prostate cancer.

### Phase 5: Developing health behavioural preventive strategies

Step 1: Assessment and Research:

- Identify Risk Factors: Understand the risk factors for prostate cancer, such as age, family history, race, and lifestyle factors.
- Gather Data: Use data collected from the baseline phases on the target population’s current behaviour, knowledge, and attitudes towards prostate cancer prevention.

Step 2: Goal Setting:

- Define Objectives: Set clear, measurable goals for the health promotion program. For example, increasing awareness about prostate cancer screening or promoting healthy lifestyle changes.

Step 3: Strategy Development:

- Educational Campaigns: Develop educational materials and campaigns to raise awareness about prostate cancer risk factors and the importance of early detection.
- Behavioral Interventions: Design interventions to promote healthy behaviors with the professional application of the selected model (Ottawa Charter & TPB) and generic activities such as regular exercise, a balanced diet, and smoking cessation.

Table 1 is an overview representation of the sub-studies within the research, linking each phase to its corresponding study objective, study design and data sources.

**Table 1:**
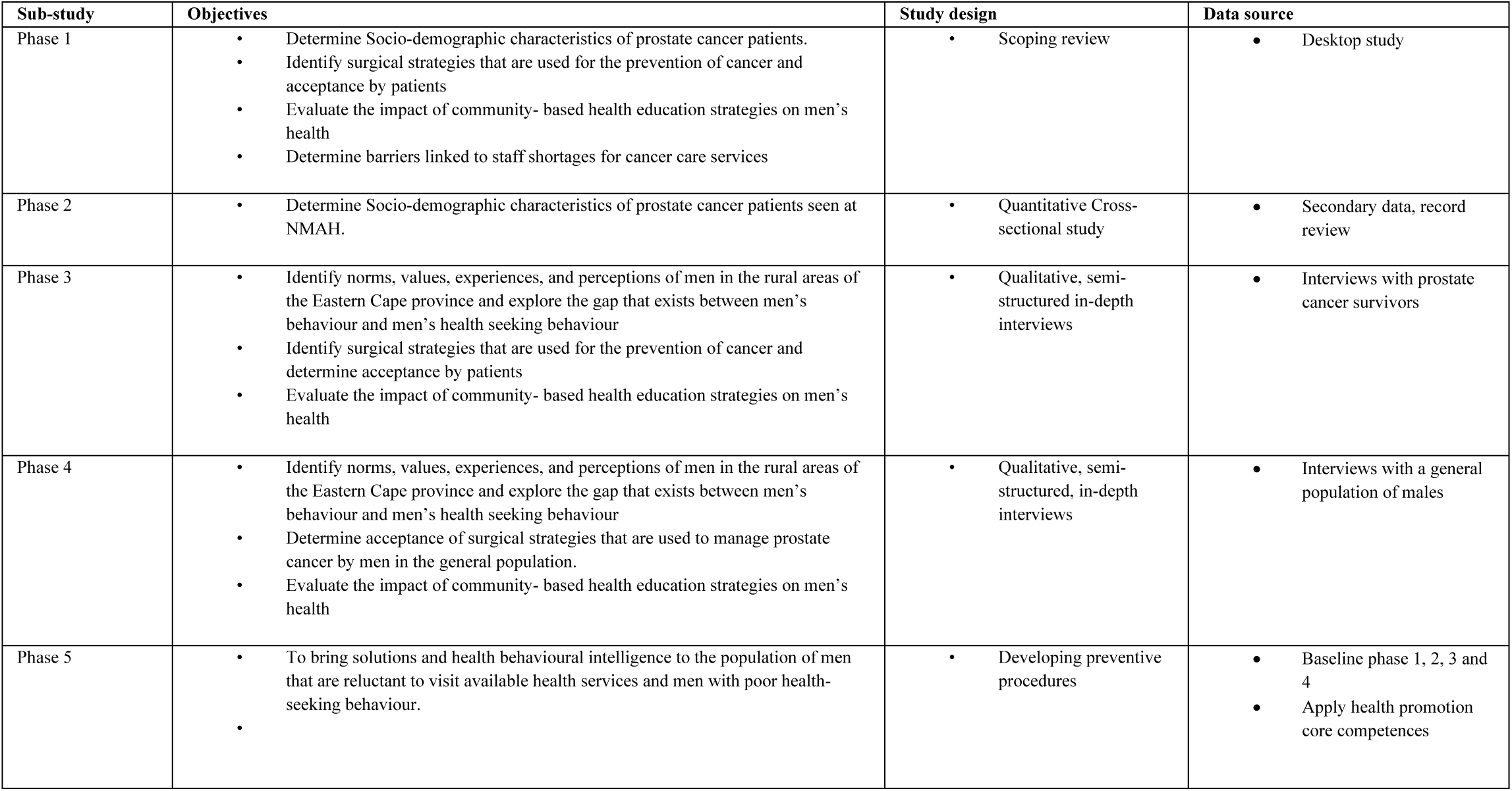
A link between each sub-study with the study’s.

| Sub-study | Objectives | Study design | Data source |
| --- | --- | --- | --- |
| Phase 1 | Determine Socio-demographic characteristics of prostate cancer patients.<br>Identify surgical strategies that are used for the prevention of cancer and acceptance by patients<br>Evaluate the impact of community- based health education strategies on men's health<br>Determine barriers linked to staff shortages for cancer care services | Scoping review | Desktop study |
| Phase 2 | Determine Socio-demographic characteristics of prostate cancer patients seen at NMAH. | Quantitative Cross-sectional study | Secondary data, record review |
| Phase 3 | Identify norms, values, experiences, and perceptions of men in the rural areas of the Eastern Cape province and explore the gap that exists between men's behaviour and men's health seeking behaviour<br>Identify surgical strategies that are used for the prevention of cancer and determine acceptance by patients<br>Evaluate the impact of community- based health education strategies on men's health | Qualitative, semi-structured in-depth interviews | Interviews with prostate cancer survivors |
| Phase 4 | Identify norms, values, experiences, and perceptions of men in the rural areas of the Eastern Cape province and explore the gap that exists between men's behaviour and men's health seeking behaviour<br>Determine acceptance of surgical strategies that are used to manage prostate cancer by men in the general population.<br>Evaluate the impact of community- based health education strategies on men's health | Qualitative, semi-structured, in-depth interviews | Interviews with a general population of males |
| Phase 5 | To bring solutions and health behavioural intelligence to the population of men that are reluctant to visit available health services and men with poor health-seeking behaviour. | Developing preventive procedures | Baseline phase 1, 2, 3 and 4<br>Apply health promotion core competences |

## Population and Sampling Population

### Inclusion Criteria

#### Phase 1: Scoping review

There has been a substantial body of research that has been conducted across the globe that focus on prostate cancer prevention and epidemiology. These studies have explored the prevalence, risk factors and socio-economic determinants of prostate cancer within different African populations. They have also examined the effectiveness of several prevention strategies and the impact of cultural beliefs on the outcome of the disease. It’s crucial to understand these factors for the development of tailored interventions and public health policies aimed at reducing the incidence and improving the early detection and treatment of prostate cancer in African settings. This is why the scoping review will focus on studies conducted in Africa. The databases that will be used are namely, ProQuest education, EBSCOhost, Science Direct and PubMed. The search term (“epidemiology” OR “incidence” OR “prevalence”) AND (“prevention” OR “intervention”) AND “prostate cancer”.

#### Phase 2: Quantitative Cross-sectional study

This sub-study will make use of secondary data of patients who were seen at Nelson Mandela Academic hospital between March 2020 and November 2021.

Sample size calculation:

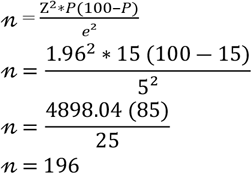

- Expected proportion is 15% of people who have been diagnosed with Prostate cancer (According to the 2019 Cancer registry, 1 in 15 men in South Africa face the lifetime risk of prostate cancer).
- Confidence level is 95%
- The maximum margin of error is 5%

According to the formula the minimum sample size is 196 patients. When allowing a 10% for non-response [sample size ÷ 0.9, (90% response)], the new targeted sample size is 217 individuals.

#### Phase 3: Qualitative, Semi-structured in-depth interviews

Patients who were diagnosed with Prostate cancer with confirmed pathology results of a prostate cancer diagnosis. Diagnosis should have been made at least six months before the date of interview to ensure that they would have had an opportunity to explore the different treatment options. The sub-study will employ a purposive sampling method, the sub-studies will also target data saturation to ensure an adequate sample size is achieved. This will allow the gathering of comprehensive insights that might not be captured if the sampling would stop at a predetermined sample size. This sampling strategy will also ensure the validity and richness of the study on cancer patients.

#### Phase 4: Qualitative, Semi-structured in-depth interviews

Persons born as males, and over 40 years of age who reside in the OR Tambo district will be interviewed in their place of residence. This sub-study will use a purposive sampling method, where researchers will sample until a point of saturation is reached. This will ensure that the data collected is rich and diverse enough to fully understand the experiences of the men who will be sampled.

### Exclusion Criteria

#### Phase 1

Scoping review

Abstracts, poster presentations and pilot studies will be excluded. Articles without English translations will also be excluded.

#### Phase 2

Quantitative Cross-sectional study

Data with missing age will be excluded as this is a key socio-demographic variable.

#### Phase 3

Qualitative, Semi-structured in-depth interviews

Patients who are medically unfit to consent will be excluded.

#### Phase 4

Qualitative, Semi-structured in-depth interviews

Males over the age of 40 who do not have the capacity to consent and those with a confirmed prostate cancer diagnosis will be excluded.

### Research Instrument

#### Phase 1

Scoping review

Covidence for the scoping review. Covidence is a software that will be used for the screening and extraction of data. A data extraction tool (Appendix A) has been designed and is informed by the literature review. The elements within the tool include; title, author, country, year of publication, aim/purpose, study design, sources of data, population/sample size, data collection tools, intervention description, results, reasons for success, key findings, conclusions, recommendations and reported gaps.

#### Phase 2 (S3 Fig)

Quantitative Cross-sectional study

The instrument for this sub-study is structured to systematically extract and analyse secondary data from medical records of male patients diagnosed with cancer and treated at the Nelson Mandela Academic Hospital’s Oncology or urology department between March 2020 and November 2021. The instrument is designed to collect key variables, including patient demographics (age, sex, and ethnicity), clinical data (type and stage of cancer, treatment modalities and outcomes), and health service utilisation (number of visits, types of interventions received, and follow-up details). The development of this instrument involved a thorough review of the hospital’s patient records system, ensuring that the variables selected are relevant for assessing the clinical journey and outcomes of cancer patients within the specified period. Standardisation of data fields was done to ensure consistency and comparability across records.

#### Phase 3 (S4 Fig)

Qualitative, Semi-structured in-depth interviews:

An interview guide has been designed for the purpose of conducting semi-structured interviews. The interview guide will have three areas of interest, a section for “opening enquiry” as well as follow-up questions. The instrument will be systematically designed to gather comprehensive data on demographic characteristics, cultural norms and values, personal experiences and perceptions, discrepancies between behaviour and health seeking practices, awareness of surgical strategies for cancer prevention, and community-based-health education. The key variables assessed by the interview guide include cultural norms and values, health-seeking behaviour, perceptions of healthcare services, acceptance of cancer prevention strategies, and the impact of community education initiatives. Audio recordings will be supplemented by field notes to preserve the exact language. The interviews will be conducted in IsiXhosa, with verbatim transcriptions. These transcriptions will undergo a process of translation and subsequent back-translation to ensure accuracy and fidelity to the original content.

#### Phase 4 (S5 Fig)

Qualitative, Semi-structured in-depth interviews:

The interview guide will be structured to comprehensively explore the norms, values, experiences and perceptions of men, with a focus on identifying the gap between their behaviour and health practices. The guide will be divided into specific sections, each targeting key areas of interest. The first section will gather demographic information, followed by an exploration of cultural norms and values that will influence men’s attitudes towards health. Subsequent sections will delve into their personal experiences and perceptions of healthcare, particularly in relation to seeking health care. Another critical area of exploration will be the acceptance of surgical strategies used to manage prostate cancer, assessing how these interventions are perceived by men in the general population. Audio recordings will be supplemented by field notes to preserve the exact language. The interviews will be conducted in IsiXhosa and therefore the transcriptions will have to be translated and back translated.

### Data quality assurance, Trustworthiness and, Validity and Reliability

#### Phase 1

Scoping review

For the Scoping Review, data quality assurance will be ensured by having two reviewers for titles, abstracts and full text screening of articles. A third reviewer will serve as a referee to resolve discrepancies throughout the process. Two reviewers will be responsible for data extraction and quality assurance. Quality assurance will use the Cochrane risk of Bias tool version 2.0 which assesses quality on five domains, namely, selection, performance, attrition, reporting and other.

#### Phase 2

Quantitative Cross-sectional study

Validity has been ensured by selecting a sample that represents the study’s objectives and the broader patient population of males that is treated at NMAH for prostate cancer. The instruments will be evaluated for the validity of the content by consulting experts in prostate cancer and public health to ensure that the tools comprehensively cover all relevant aspects of prostate cancer. Reliability will be addressed by adopting rigorous data handling procedures and thorough cleaning of the data.

#### Phase 3 and 4

Qualitative, Semi-structured in-depth interviews

To ensure rigor several strategies will have to be employed to enhance trustworthiness and credibility of the findings following principles outlined by Ritchie [16]. Namely:

1. Trustworthiness: This will involve clearly documenting the research process and ensuring transparency in data collection and analysis
2. Audit trail: an audit trail will be maintained throughout the study, this will provide a detailed account of all research decisions, data collection, and data analysis procedures. This documentation will allow others to follow the research process and assess the consistency and dependability of the findings.
3. Peer checking: to enhance credibility and reduce researcher bias, peer checking will be employed. Colleagues with expertise in qualitative research and prostate cancer will review the data and preliminary findings to provide feedback and ensure that the interpretations are sound and unbiased.
4. Member checking: After data analysis, member checking will be conducted by sharing preliminary findings with a subset of participants. This will allow participants to confirm the accuracy of the interpretations and provide additional insights, this will enhance the credibility of the results.
5. Triangulation: By comparing data from different sources such as interviews, field notes and secondary data the results will be corroborated. Thus, ensuring that the findings are consistent and reliable across multiple data points.

### Reflexivity

Customary to qualitative study research, this study will disclose reflexivity of the researcher as a black, South African female of the amaXhosa tribe who is married with male children, I have intricate knowledge of the cultural practices of the community. The researcher is a health promoter and a researcher with a Master of Public Health qualification who is undertaking this research as part of her doctoral studies. In this culture where participants will be interviewed, it is difficult to openly discuss male sexual organs and male sexuality when conversing with males or elders. The Researcher will therefore have to overcome the cultural barriers and stereotypes of being perceived to be wanting to be treated differently because of her education and perceived socio-economic status when undertaking this research. Having been raised in rural areas of the Eastern Cape, the researcher possesses an understanding of the social cues specific to amaXhosa men.

### Data Management and Analysis

#### Phase 1

Scoping Review

A narrative synthesis of articles will be undertaken. Tables and Figures will be used where appropriate. A Prisma diagram will be used to summarise the articles searched, included and those excluded with reasons for exclusion.

#### Phase 2

Quantitative Cross-sectional study

Data will be captured and coded in Microsoft excel 2013 (Microsoft corporation, Seattle, USA). The data will then be exported to STATA 18 for analyses. The data will be summarised through the use of descriptive tables, graphs and means (with standard deviation and range values) or medians (with the interquartile range) depending on the normality of distribution of numerical variables. Tests that will be used for numerical variables are;

- T-test or Wilcoxon rank-sum test: to compare the mean or median ages of patients from different referral facilities and ascertain if these ages differ by characteristics (e.g., early diagnosis vs late diagnosis)
- ANOVA or Kruskal Wallis test: will be used to compare continuous outcomes (means or medians) across multiple groups to identify significant differences in ages related to cancer stages or treatments.

For Categorical Variables:

- Logistic regression: This test will help identify factors that influence the probability of patients having a successful treatment outcome.
- Chi-square test: will be used to determine if there is a significant relationship between variables such as treatment and patient outcomes

#### Phase 3 and 4

Qualitative, Semi-structured in-depth interviews

Transcribed data will be exported to NVIVO 14 for analysis. Thematic coding will be used to identify themes within the data. Inductive analyses will be used to help develop themes which emerge from the data. Themes will be derived using a six steps approach of familiarisation, coding, theme development, review of themes, defining of themes and reporting (cite literature on the six steps approach). Rich data will be represented by having multiple direct quotes from participants. Data will be summarised using graphs and Tables.

### Patient and public Involvement

The research questions were informed by gaps identified during the review of literature that has been published over time on Prostate Cancer. Findings from systematic reviews, scoping reviews and meta-analyses that focus on the lived experiences of survivors and health-seeking behaviors of men in rural areas around Africa also informed the research questions. Furthermore, the research objectives were developed after reviewing the literature, identifying knowledge gaps, and noting that some systematic reviews on prostate cancer prevention yielded inconclusive results. Additionally, these reviews highlighted stakeholder priorities, which further informed the focus of the study.

In terms of patient involvement, input will be sought from patients to ensure the study design is culturally sensitive and that it aligns with their lived experiences.

Awareness, screening, support, early access, acceptability will be reflected in the outcome measures. Survivors, community leaders, the Oncology unit at Nelson Mandela Academic Hospital will assist in reaching relevant participants effectively.

### Ethics and Dissemination

This study will abide by the four ethical principles of autonomy, beneficence, non-maleficence and justice and approved by Walter Sisulu University Health Sciences Research Ethics Committee for ethics approval. For sub-study 2, where secondary data will be used, the need for consent is waivered by the anonymisation and de-identification of data. Retrospective tracking of participants is therefore impossible. There will be no personal information that could be used to re-identify individuals. These measurers will be taken to protect the data and safeguard patient information. For the other sub-studies where, primary data will be collected, informed consent will be obtained from participants before data collection. Confidentiality will be maintained by anonymizing participants through unique identifiers, and all personal information will be de-identified prior to analysis. Explicit consent for recording will be obtained, and participants will be assured that their responses will remain confidential. Data will be stored securely and will be password protected. Only the Principal investigator will have access to the data. Participants will be informed of their right to withdraw at any stage without repercussions. Upon receipt of ethics clearance, the study was submitted to the Eastern Cape Provincial Health Research Committee for research access approval and was approval granted [EC_202503_009].

The findings of this study will be disseminated through academic publications and conferences to ensure the results reach the relevant stakeholders and contribute to knowledge in the prevention of prostate cancer.

### Current status and timeline of the study

Ethics approval from Walter Sisulu University Health Science Research Ethics Committee obtained with approval number (WSU HREC 014/2025). Quantitative collection is still planned, the recruitment period begins on the 30^th^ of May 2025. Qualitative interviews have not yet commenced.

The study is structured with a clear timeline. Participant recruitment for qualitative interviews is expected to be completed by August 2025. Data collection encompassing both qualitative interviews and the scoping review is scheduled to conclude by December 2025. Data cleaning and analysis will commence immediately afterwards. Results are anticipated to be finalized by June 2026. The final write-up and dissemination of findings will follow, with the study expected to be completed by mid 2026.

## Discussions

### Strengths and Limitations of the study

- The triangulation of methods enhances the credibility and depth of findings on prostate cancer
- The study will capture rich, detailed personal experiences and perceptions about prostate cancer in low resource settings
- Flexibility allows probing for deeper understanding of survivors’ beliefs and challenges
- Participants in study area are largely illiterate, thus requiring translation of tool into local language. This introduces a bias as some body parts are not well defined in the local language.

### Amendments to the study

Should there be any modifications required, the principal investigator will review the changes and provide justification. Amendments will be submitted to the Walter Sisulu University Health Sciences Research Ethics Committee (WSUHREC) for approval before implementation

## Data Availability

No datasets were generated or analysed during the current study. All relevant data from this study will be made available upon study completion. Deidentified research data will be made publicly available when the study is completed and published.

## Authors’ Contributions

Mrs. Xolelwa Ntlongweni – Principal Investigator, responsible for conceptualizing the work by ensuring that the research question, study design and methodology align with the objectives. Integrated feedback from all supervisors to ensure the proposal meets all academic and ethical standards.

Prof. S.A. Mabunda – Mentor and Supervisor, responsible for academic support and mentorship, reviewed drafts of the research proposal and ensured that the work maintains academic rigor and quality.

Dr. S.C. Nomatshila – Co-supervisor; Gave additional academic support, supported in problem solving by offering alternative perspectives to overcome obstacles.

Prof. W.W. Chitha – Co-supervisor; Is responsible for ensuring professional growth. Ensuring there are resources that are allocated efficiently to facilitate the research process.

## Acknowledgements

The authors would like to express their gratitude and appreciation to the Walter Sisulu University Health Sciences Research Ethics and Scientific Review committee for their guidance. A special thanks to the department of Public Health housed within the Faculty of health Sciences for providing support.

## Supporting Information

S1 Fig. Map of OR Tambo District. Provides contextual background for the study site, showing the geographical layout of the OR Tambo district.

S2 Fig. Data extraction tool for scoping review sub study 1. The tool will be used to extract key information from selected studies.

S3 Fig. Data extraction tool for sub study 2. The tool will be used for extracting patient dada from medical records.

S4 Fig. Interview guide for prostate cancer survivors. This guide includes key thematic areas to explore survivors’ lived experiences, perceptions and challenges related to diagnosis, treatment and healthcare access.

S5 Fig. Interview guide for the general population of men. This guide will be used to explore knowledge, attitudes and health-seeking behaviour of men in the general population regarding prostate cancer. The guide includes key thematic areas such as assess awareness, perceptions and potential barriers to screening and treatment.

S6 Fig. Patient Consent form. This form will be used in the study to ensure that there is informed participation. The form outlines the purpose of the research, rights of the participants, confidentiality measurers and consent for data collection.

S7 Fig. Research logistics and Timelines. This shows the research logistics and projected timelines outlining key activities, responsible personnel and timeframes for completion.

